# Thermal variability and the geography of optimal temperature for child survival: childhood respiratory-infection mortality in 171 countries: a systematic analysis of the Global Burden of Disease Study 2023 and the C-LSAT high-resolution climate dataset

**DOI:** 10.64898/2026.08.31.26361864

**Authors:** Deze Li, Song Sun, Hao Chen, Xiaotong Wang, Chen Shen

**Author notes:** Correspondence to: Chen Shen, Beijing Pediatric Research Institute, Beijing Children’s Hospital, Capital Medical University, Beijing, China.

## Abstract

**Background:** In adults, cold-attributable mortality exceeds heat-attributable mortality roughly 17-fold. Child-specific evidence has begun to emerge only recently—a nationwide Brazilian case-crossover study located the minimum mortality temperature (MMT) for under-five deaths, and a 56-country survey-based analysis linked monthly temperature anomalies to under-five mortality—but no multi-country, climate-zone-resolved estimate of the childhood respiratory-infection MMT exists, and whether temperature variability is independently associated with childhood respiratory mortality at the global scale is unknown. We quantified both.

**Methods:** We combined Global Burden of Disease 2023 mortality estimates—lower respiratory infection (LRI) deaths at ages 0–19 years and asthma deaths at ages 0–24 years, 171 countries, 1990–2023—with 0.5° monthly land temperature and diurnal temperature range (DTR) fields from C-LSAT/C-LDTR (1901–2023). Four exposure dimensions (annual mean, DTR, seasonal amplitude, interannual variability) entered two-way fixed-effects models with Driscoll–Kraay standard errors. A quadratic term in mean temperature located the MMT, with percentile confidence intervals from a 300-replication country-cluster bootstrap. Future-exposure leads, country-level detrending, and permutation tests assessed contemporaneous causality, applied to both the linear coefficients and the quadratic term generating the MMT; national pneumococcal conjugate vaccine (PCV3) coverage and ambient PM2.5 exposure series were added as time-varying mechanistic covariates. Country-level exposure changes for 2041–2060 were additionally projected under three CMIP6 SSP scenarios (WorldClim v2.1 baseline; MRI-ESM2-0) using a matched-cell delta method.

**Results:** The childhood LRI MMT was 17.1 °C (95% CI 14.7–19.8), the 36th percentile of the annual-temperature distribution; zone estimates were 24.7 °C in tropical and 15.8 °C in subtropical countries, with weak temperate and no subarctic identification. The quadratic term underpinning the MMT, however, failed both falsification checks—future temperatures reproduced the U-shape and country-level detrending erased it—so these MMT values describe a trend-level geographic pattern of the annual construct rather than a contemporaneous dose– response. Interannual temperature variability was positively associated with LRI (+0.278, 95% CI 0.102–0.454; p = 0.002) and asthma mortality (+0.836, 95% CI 0.447–1.226; p = 2.6 × 10−5) per 1 °C, but future-exposure models returned nearly identical significant coefficients and detrending erased significance, supporting only a trend-level association; adjustment for national PCV3 coverage and PM2.5 exposure left these estimates essentially unchanged. Annual mean temperature was likewise inversely associated with both outcomes at the trend level; DTR and seasonal amplitude showed no independent within-country effects. Under all three scenarios, country-mean annual temperature is projected to rise by +1.6 to +2.4 °C by 2041–2060, with seasonal amplitude widening in 77–81% of the 196 projection polygons (a climate-raster set wider than the 171-country mortality panel).

**Conclusions:** This study provides the first multi-country, climate-zone-resolved geography of the optimal temperature for childhood respiratory survival, spanning 171 countries; because the underlying quadratic association is trend-level, the estimates are directional. The observed variability–mortality associations are trend-level signals rather than contemporaneous causal evidence; daily-scale, child-specific designs are required to determine whether short-term thermal variability affects paediatric respiratory mortality.

## 1. Introduction

Non-optimal ambient temperature is an established cause of premature death. In a 13-country analysis, 7.71% of mortality was attributable to temperature, with cold accounting for 7.29% against 0.42% for heat—an approximately 17-fold difference ^1^. Extending such estimates globally, an average of 5,083,173 deaths per year (9.43% of global mortality) were associated with non-optimal temperatures during 2000–2019 ^2^. The temperature at which mortality is lowest—the minimum mortality temperature (MMT)—is not constant across climates: it falls near the 60th percentile of the local temperature distribution in tropical regions but at the 80th–90th percentile in temperate zones, indicating substantial human adaptation to local climate ^13^.

Beyond the thermal level, short-term temperature variability also affects adult health. Diurnal temperature range (DTR) has been linked to mortality in single-city and multi-city studies—for example, a 1 °C increase in DTR was associated with a 1.29% increase in respiratory mortality on cold days in Shanghai ^4^—and multi-country analyses of temperature variability more broadly ^5^ estimate that 2.5% of deaths are attributable to DTR ^6^. Global three-stage modelling studies have since quantified the mortality burdens of short-term temperature variability ^7^, its intra-day versus inter-day components ^8^, heatwaves ^9^, and cold spells ^10^. Systematic reviews confirm generally consistent harmful effects of large DTR on cardiovascular and respiratory outcomes ^11^. This adult literature rests on mature daily time-series methods, notably distributed lag non-linear models (DLNMs), which flexibly capture non-linear and delayed temperature–mortality associations ^12,13^.

Children, by contrast, remain largely absent from this evidence base. A 2012 systematic review concluded that infants under one year are the most vulnerable to heat-related death and that temperature affects children mainly through infectious diseases, including respiratory infections, while explicitly identifying the effect of temperature variability on children as a research gap ^14^. The subsequent paediatric literature has been narrow: DTR has been associated with childhood asthma only through emergency-department visits (a 5 °C increase in DTR with a 31% [95% CI 11–58%] rise in presentations) ^15^, and extreme temperature has been linked to paediatric respiratory outpatient visits in a single Chinese city ^16^. Exposure is also intensifying: infants worldwide experienced an average of 13.8 days of heatwave in 2023, 8.2 days more than the 1986–2005 baseline ^17^. Two 2025 studies have begun to close the child-mortality gap at finer temporal scales: a space-time-stratified case-crossover analysis of 1,061,229 under-five deaths in Brazil estimated a U-shaped temperature–mortality relationship against an MMT reference— respiratory deaths being associated only with heat ^18^—and a sibling-matched analysis of 1,745,132 live births across 56 low– and middle-income countries linked monthly temperature anomalies to under-five mortality with pronounced climate-zone heterogeneity ^19^. What remains missing is a multi-country estimate that spans all climate zones, is specific to respiratory causes, and decomposes the dimensions of thermal variability: no multi-country, climate-zone-resolved MMT estimate exists for childhood respiratory mortality, and interannual temperature variability has not been examined against child mortality at the global scale.

We address these gaps in three ways. First, combining Global Burden of Disease (GBD) 2023 mortality estimates for ages 0–19 years (lower respiratory infections) and 0–24 years (asthma) with 0.5° gridded climate data ^20^, we derive the first multi-country, climate-zone-resolved geography of the childhood respiratory-infection MMT across 171 countries over 1990–2023—an annual ecological complement to the daily and monthly individual-level designs of recent national and multi-country studies ^18,19^. Second, we decompose four dimensions of thermal exposure—annual mean temperature, DTR, seasonal amplitude, and interannual variability— estimating their independent associations while controlling for the mean level. Third, because annual ecological panels can confound common trends with contemporaneous effects, we embed a pre-specified falsification framework (future-exposure leads, within-country detrending, and permutation tests) that explicitly separates trend-level associations from same-year causal effects, and we report both findings and failed falsifications.

## 2. Methods

### 2.1 Data sources

Mortality estimates were obtained from the Global Burden of Disease Study 2023 (GBD 2023), which produces internally consistent cause-specific death estimates for 204 countries and territories from 1990 to 2023 ^21,22,23^. We newly extracted national-level deaths from lower respiratory infections (LRI; GBD cause 322) for children aged 0–19 years, computed as the sum of the four GBD age groups <5, 5–9, 10–14, and 15–19 years, and deaths from asthma (GBD cause 515) for ages 0–24 years, via the GBD Results Tool ^24^. Asthma served as a second respiratory-related outcome rather than a placebo.

Temperature data were derived from the High-Resolution China global Land Surface Air Temperature version 1 (C-LSAT HRv1) and the paired diurnal temperature range product (C-LDTR HRv1): monthly 0.5° × 0.5° gridded land fields for 1901–2023 ^20^. Absolute monthly temperature and diurnal temperature range (DTR) were reconstructed as the 1961–1990 monthly climatology plus the monthly anomaly. Grid cells were assigned to countries using Natural Earth 50 m boundaries, and country-year values were computed as cosine-of-latitude area-weighted means over valid land cells; a cell-year required 12 finite months. Countries without any polygon-covered land cell (mostly small islands) had been assigned their nearest land data cell (nearest_cell, n = 55 in the source file). Of the 204 GBD countries, 201 matched the climate file; the main panel excludes the 30 nearest_cell small-island states intersecting GBD estimates, yielding 171 polygon-masked countries over 1990–2023 (5,814 country-years). Six polygon climate territories have no GBD estimates and three GBD countries (France, Norway, Tokelau) lack climate records, so 171 is the structural maximum of the intersection. A sensitivity analysis re-includes the 30 small-island states (201 countries).

### 2.2 Exposure construction

Four country-year temperature indicators were constructed for 1990–2023: mean_temp, the area-weighted annual mean land surface air temperature (°C); dtr_mean, the area-weighted annual mean DTR (°C); season_amp, the warmest-month minus coldest-month difference of the 12 area-weighted monthly mean temperatures (°C); and interannual_sd, the standard deviation of annual mean temperature over a trailing 10-year window (year y uses years y−9 to y). These metrics separate the thermal level from within-day, within-year, and between-year variability.

### 2.3 Outcomes and rates

Outcomes were national counts of LRI deaths at ages 0–19 years and asthma deaths at ages 0–24 years in each country and year. Mortality rates were computed per 100,000 using the GLOBOCAN 2022 population aged 0–19 years as a fixed denominator for all years. This fixed denominator preserves the temporal structure of the GBD death estimates but ignores population growth and ageing, a limitation we acknowledge; because the denominator is a time-invariant constant within each country, it is fully absorbed by country fixed effects and does not affect fixed-effect coefficients. Four countries (Greenland, Saint Vincent and the Grenadines, Taiwan, United States Virgin Islands) lack this denominator; they are retained in fixed-effect models but excluded from descriptive rate statistics. The asthma rate uses the 0–19 denominator although its outcome spans 0–24 years, slightly inflating rate levels without affecting fixed-effect coefficients.

### 2.4 Main model

The primary specification regressed the log mortality rate on the four temperature indicators with two-way fixed effects: log(rate) ∼ mean_temp + dtr_mean + season_amp + interannual_sd, with country and year fixed effects. Country fixed effects absorb all time-invariant heterogeneity (geography, latitude, baseline development); time-varying trajectories common to all countries are absorbed by year fixed effects. Driscoll–Kraay standard errors (bandwidth lag = 3) are robust to heteroskedasticity, autocorrelation, and cross-sectional dependence. We report log-rate coefficients with exponentiated percentage changes in the mortality rate per 1 °C change in exposure. The year-fixed-effect covariate model (154 countries; no country fixed effects, so coefficients reflect both cross-sectional and temporal variation) additionally includes log GDP per capita, PM2.5 exposure and urban population share. Because air pollution is time-varying and may confound trend-level temperature associations, we additionally extracted the GBD 2023 national annual exposure series for ambient particulate matter pollution — the age-standardized summary exposure value (SEV), a modelled 0–100 exposure index produced within the GBD risk-factor framework ^23^ — for all 204 countries over 1990–2023, and re-fitted the main two-way fixed-effects model with this PM2.5 time series as a covariate, with and without GBD-super-region-specific linear year trends. Because childhood vaccination is the most proximal mechanistic driver of declining LRI mortality and its roll-out was staggered across countries, we additionally merged national annual third-dose pneumococcal conjugate vaccine (PCV3) coverage from the WHO/UNICEF estimates of national immunization coverage (WUENIC, 2025 revision) ^25^ into the panel by country and year—years before 2008, preceding vaccine availability and the WUENIC series, were set to zero—and re-fitted the main model with PCV3 coverage as a time-varying covariate, on the full panel and restricted to 2008–2023. Year fixed effects absorb the global COVID-19 shock; country-differential pandemic impacts are not separately modelled (GBD 2023 allocates COVID-19 deaths to a dedicated cause, so LRI envelopes are not directly inflated by the pandemic’s cause-of-death reassignment).

### 2.5 Minimum mortality temperature

To estimate the minimum mortality temperature (MMT), we added a quadratic term for mean_temp to the main model and computed the turning point as −b1/(2 × b2), with b2 > 0 indicating a U-shaped association ^3^. Models were fitted overall and by climate zone (tropical, subtropical, temperate, subarctic), with percentile confidence intervals from a 300-replication bootstrap resampling countries. Because our exposure is the country-level annual mean temperature rather than the daily temperature distributions used in adult MMT studies, comparisons with adult estimates are directional only ^1^. Temporal aggregation from daily to weekly, monthly, or annual constructs is known to bias both the tails and the estimated MMT of temperature–mortality curves ^26^, so our MMT is explicitly an annual-construct estimate. And because the MMT is derived from the quadratic coefficients, the future-exposure and detrending checks of Section 2.6 were additionally applied to the quadratic specification itself, reporting the stability of the quadratic coefficient b2 and of the implied MMT.

### 2.6 Falsification suite

Given the ecological design, we pre-specified four checks, applied to the main specification and, for the first two, to the quadratic MMT specification. First, future-exposure tests replaced exposures with their t + 1 and t + 2 values; future-temperature coefficients resembling contemporaneous ones would indicate trend-level co-movement rather than a same-year effect. Second, country-specific linear detrending separated trend-level from year-to-year variation. Third, a permutation test shuffled years within each country 200 times to assess whether associations depend on true temporal ordering. Fourth, residual spatial autocorrelation was assessed with Moran’s I (k-nearest-neighbour weights, k = 5) on country-mean residuals.

### 2.7 Reporting, ethics, and software

. The study used publicly available, de-identified, aggregated estimates and was exempt from ethics review. Analyses were conducted in Python (pandas, statsmodels, spreg); all random procedures used fixed seeds, and the analysis code is available from the corresponding author upon reasonable request (see Availability of data and materials).

### 2.8 Climate-scenario exposure projection to the 2050s

As forward-looking context, we projected country-level changes in three of the four thermal-exposure indicators under CMIP6 climate scenarios. Monthly minimum and maximum temperature fields were taken from WorldClim version 2.1 (1970–2000 baseline climatology, 10 arc-minute resolution)^28^ and from the corresponding CMIP6-downscaled projections for 2041– 2060 under SSP1-2.6, SSP2-4.5 and SSP5-8.5, produced with a single general circulation model (MRI-ESM2-0) and distributed through the WorldClim CMIP6 archive. Country polygons (Natural Earth 10 m admin-0) were rasterised onto the 10 arc-minute grid and, for each country and scenario, baseline and future means were computed over the same grid cells — those valid in the future land mask — so that every estimate is a matched-cell change. Countries whose polygons required buffer assignment to intersect any future land cell (mostly small island states) were omitted, leaving 196 Natural Earth admin-0 polygons (including Antarctica, dependent territories and disputed units; this projection set is not identical to the 171-country mortality panel). Per country, we computed the change in annual mean temperature (mean of monthly (tmin + tmax)/2), in mean diurnal temperature range (mean of monthly tmax − tmin) and in seasonal amplitude (warmest minus coldest monthly mean temperature); summaries are equal-weighted across countries. Interannual variability cannot be derived from 20-year climatological means and is therefore not projected. Given the trend-level nature of the mortality associations (Section 3.4), these projections characterise future exposure only; no mortality or burden projection is attempted.

## 3. Results

### 3.1 Analytical panel and descriptive patterns

As internal validation, summed 2023 lower respiratory infection (LRI) deaths across the full 204-country CSV (711,228.5) reproduced the GBD 2023 global estimate (711,228) to within 0.001%; the 171-country analytical panel sums to 711,010.8, 0.03% below the global value, the difference being the 30 island nearest-cell countries and the 3 countries without climate correspondence (France, Norway, Tokelau). stable seasonal cycle (Figure 1)

**Figure 1.**
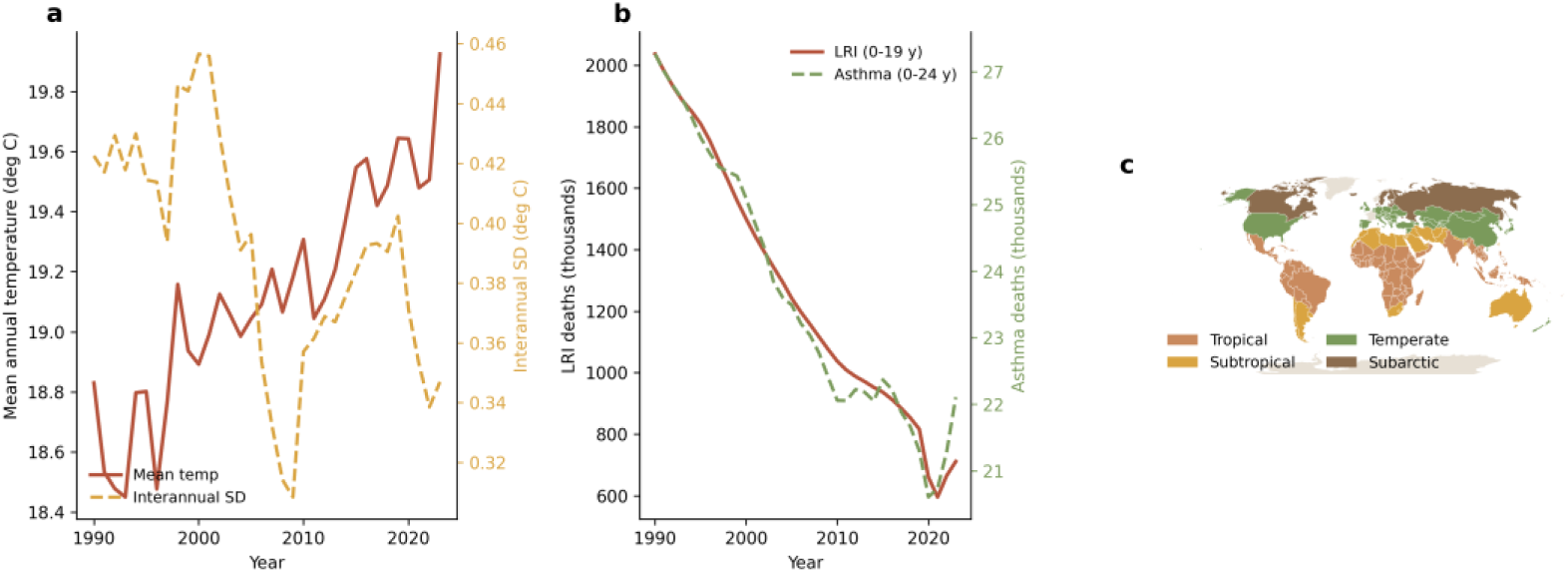
(a) Global mean annual temperature (solid) and interannual temperature variability (dashed), 171-country main panel, 1990–2023; (b) global childhood deaths from lower respiratory infections (LRI, ages 0–19) and asthma (ages 0–24), 1990–2023; (c) data coverage map: climate-zone assignment of the 171 panel countries (Robinson projection; non-panel countries in grey).

**Table 1.** Structure of the analytical panel, 171 countries, 1990–2023.

| **Item** | **LRI (ages 0–19)** | **Asthma (ages 0–24)** |
|---|---|---|
| GBD 2023 countries/territories | 204 | 204 |
| Matched to climate data | 201 | 201 |
| Main panel (polygon-masked) | 171 | 171 |
| Nearest-cell small island states (sensitivity only) | 30 | 30 |
| Unmatched to climate file | France, Norway, Tokelau | France, Norway, Tokelau |
| Country-year observations (main panel) | 5,814 | 5,814 |
| Study period | 1990–2023 | 1990–2023 |
| Missing fixed 2022 denominator (absorbed by country FE) | 4 countries | 4 countries |
| Missing covariates (GDP per capita, PM2.5, urban population share) | 17 countries | 17 countries |

### 3.2 Main effects of temperature metrics on childhood respiratory mortality

Figure 1 and Table 2 present the two-way fixed-effects estimates with Driscoll–Kraay standard errors; with log-transformed outcomes, coefficients are log-rate effects, which we also report as exponentiated percentage changes per 1 °C. Annual mean temperature was inversely associated with both outcomes: −0.0721 (95% CI −0.1239 to −0.0203; p = 0.0063) for LRI mortality at ages 0–19 and −0.1102 (95% CI −0.1829 to −0.0374; p = 0.0030) for asthma mortality at ages 0–24 (exponentiated: approximately 7% and 10% lower rates per 1 °C). Interannual variability was positively associated with both: +0.2780 (95% CI 0.1020 to 0.4539; p = 0.0020) for LRI and +0.8363 (95% CI 0.4469 to 1.2257; p = 2.6 × 10−5) for asthma (exponentiated: +32% for LRI and +131% for asthma per 1 °C). Neither DTR (LRI: 0.0202, 95% CI −0.0474 to 0.0878, p = 0.5575; asthma: 0.0249, 95% CI −0.0712 to 0.1210, p = 0.6116) nor seasonal amplitude (LRI: −0.0085, 95% CI −0.0214 to 0.0043, p = 0.1938; asthma: −0.0058, 95% CI −0.0255 to 0.0140, p = 0.5670) showed a significant independent association. Within-country R² was 0.097 (LRI) and 0.166 (asthma).

**Table 2.**
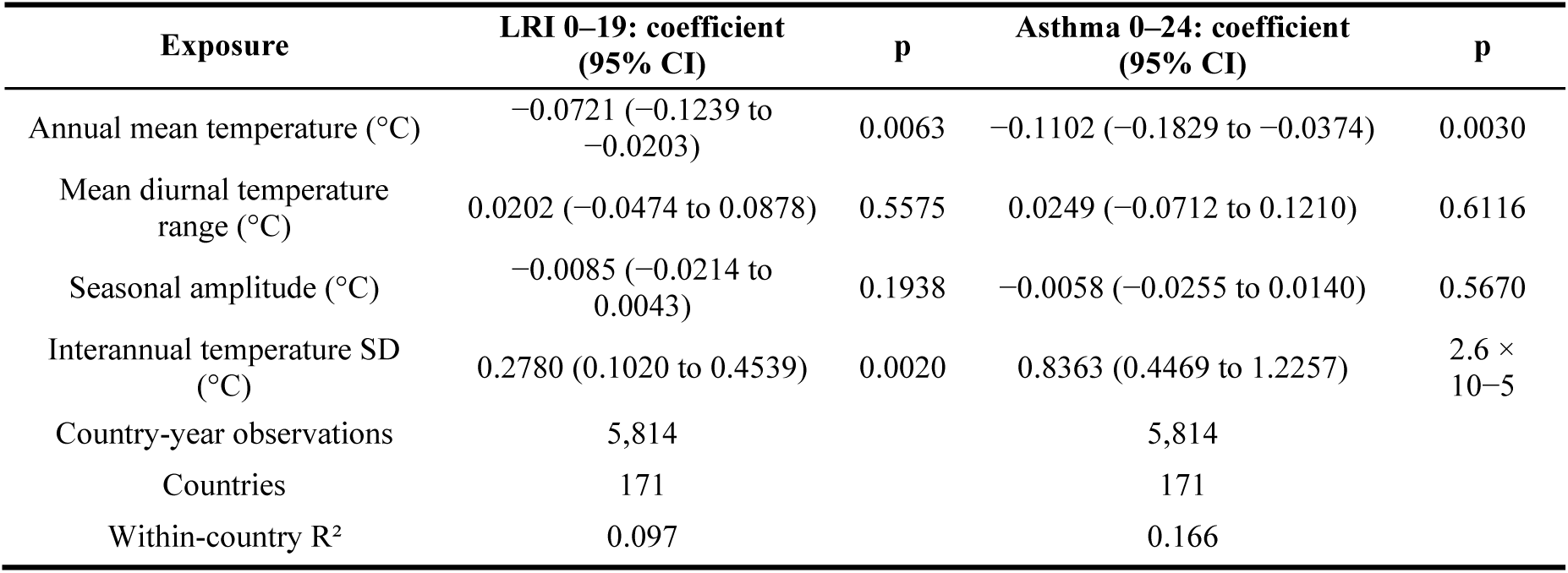
Two-way fixed-effects estimates of temperature metrics against log childhood respiratory mortality, 171 countries, 1990–2023 (Driscoll–Kraay standard errors, bandwidth 3)

| **Exposure** | **LRI 0–19: coefficient (95% CI)** | **p** | **Asthma 0–24: coefficient (95% CI)** | **p** |
|---|---|---|---|---|
| Annual mean temperature (°C) | −0.0721 (−0.1239 to −0.0203) | 0.0063 | −0.1102 (−0.1829 to −0.0374) | 0.0030 |
| Mean diurnal temperature range (°C) | 0.0202 (−0.0474 to 0.0878) | 0.5575 | 0.0249 (−0.0712 to 0.1210) | 0.6116 |
| Seasonal amplitude (°C) | −0.0085 (−0.0214 to 0.0043) | 0.1938 | −0.0058 (−0.0255 to 0.0140) | 0.5670 |
| Interannual temperature SD (°C) | 0.2780 (0.1020 to 0.4539) | 0.0020 | 0.8363 (0.4469 to 1.2257) | 2.6 × 10−5 |
| Country-year observations | 5,814 |  | 5,814 |  |
| Countries | 171 |  | 171 |  |
| Within-country R² | 0.097 |  | 0.166 |  |

Figure 1 highlights two patterns. First, the significant effects run in consistent directions across outcomes: warmer mean conditions track lower mortality, whereas larger interannual fluctuations track higher mortality. Second, the asthma estimates exceed the LRI estimates in magnitude, with wider intervals, while the DTR and seasonal-amplitude nulls are tightly centred near zero, arguing against a substantial independent within-country effect of either metric at the annual scale. As Section 3.4 shows, the significant associations are trend-level associations, not contemporaneous causal evidence.

### 3.3 Minimum mortality temperature in children

Adding a quadratic term in annual mean temperature identified a U-shaped temperature–mortality relationship for childhood LRI deaths (Figure 2; Table 3). The overall minimum mortality temperature (MMT) was 17.1 °C (95% CI 14.7 to 19.8; 300 of 300 valid bootstrap replicates), at the 36th percentile of the pooled annual-temperature distribution. By climate zone, the MMT was 24.7 °C (95% CI 18.5 to 27.0) in tropical countries (86 countries; 32nd percentile) and 15.8 °C (95% CI 3.1 to 20.1) in subtropical countries (29 countries; 21st percentile). Identification was weak elsewhere: the temperate estimate (46 countries) was 25.5 °C (95% CI 8.1 to 28.3), sitting at the upper boundary of the observed distribution (100th percentile) with only 181 of 300 valid replicates, and in the nine subarctic countries the quadratic coefficient was negative (an inverted U), so no minimum was identified. The temperate and subarctic results should be read as uninformative rather than as evidence of a high-latitude optimum. Moreover, as Section 3.4 shows, the quadratic form underpinning these estimates is itself trend-level: the MMT geography should be read as a directional pattern of the annual construct, not as a contemporaneous dose– response relationship.

**Figure 2.**
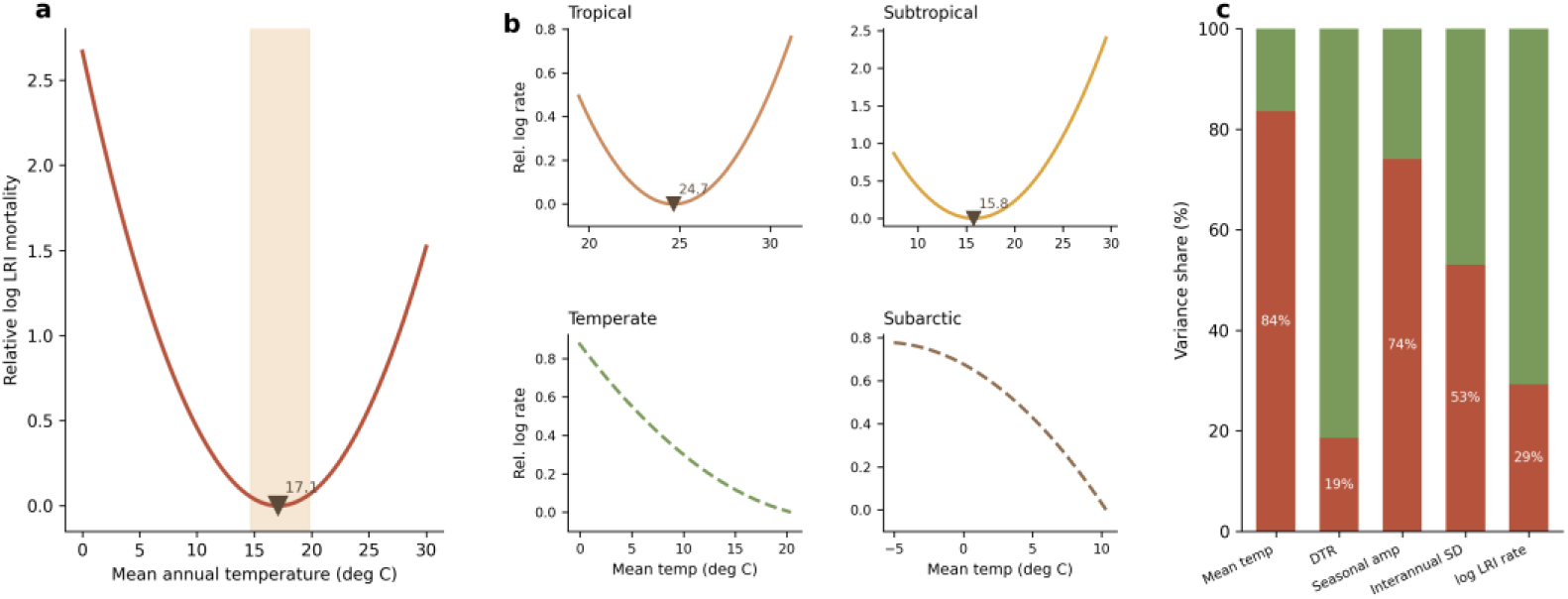
(a) Overall quadratic temperature–mortality curve for LRI with the minimum mortality temperature (MMT = 17.1 °C; bootstrap 95% CI 14.7–19.8 shaded); (b) zone-specific quadratic curves (small multiples; solid = significant quadratic term, dashed = non-significant or no finite MMT; MMT marked where inside the zone’s observed range); (c) variance decomposition of the four thermal metrics and log LRI mortality into between-belt and within-belt shares.

**Table 3.** Minimum mortality temperature (MMT) for childhood LRI mortality (ages 0–19) by climate zone, quadratic fixed-effects models with country-cluster bootstrap (300 replicates) Note: One country (34 country-years) could not be assigned a climate zone and is excluded from the zone rows.

| **Climate zone** | **Countries** | **Country-years** | **MMT (°C)** | **95% CI (°C)** | **Percentile of zone temperature distribution** | **Valid bootstrap replicates** |
|---|---|---|---|---|---|---|
| Overall | 171 | 5,814 | 17.1 | 14.7 to 19.8 | 36th | 300 |
| Tropical | 86 | 2,924 | 24.7 | 18.5 to 27.0 | 32nd | 289 |
| Subtropical | 29 | 986 | 15.8 | 3.1 to 20.1 | 21st | 285 |
| Temperate | 46 | 1,564 | 25.5 | 8.1 to 28.3 | 100th (upper boundary; weak identification) | 181 |
| Subarctic | 9 | 306 | Not identified (no U-shaped minimum) | — | — | 84 |

Two features of Figure 2 deserve emphasis. First, where identification is adequate, the child MMT falls well below the midpoint of the local temperature distribution (21st–36th percentiles): within this annual-resolution construct, minimum childhood LRI mortality occurs at temperatures cooler than the typical local climate, with mortality rising steeply on the warm side of the optimum in tropical settings. Second, these percentiles lie far below the 60th–90th percentile values reported in adult daily-temperature studies; because our exposure is the annual mean rather than the daily temperature distribution, the constructs are not directly commensurable, and the contrast warrants caution.

### 3.4 Falsification and sensitivity analyses

The falsification suite (Figure 3) qualified these findings. First, lead-exposure models regressing mortality on temperatures one or two years ahead returned coefficients nearly identical to the contemporaneous model — for LRI, the lead coefficients were 0.308 (t + 1) and 0.337 (t + 2), both p < 0.001, while the contemporaneous coefficient was 0.278 (p = 0.002). Future temperatures cannot cause past deaths, so this test fails: the associations reflect slow-moving co-trends, and we characterise them throughout as trend-level associations, not contemporaneous causal evidence. Consistently, country-specific linear detrending attenuated all four coefficients towards zero and eliminated significance for the variability and mean-temperature terms (the seasonal-amplitude term remained marginal at p = 0.045; LRI interannual variability: 0.278 to −0.078).

**Figure 3.**
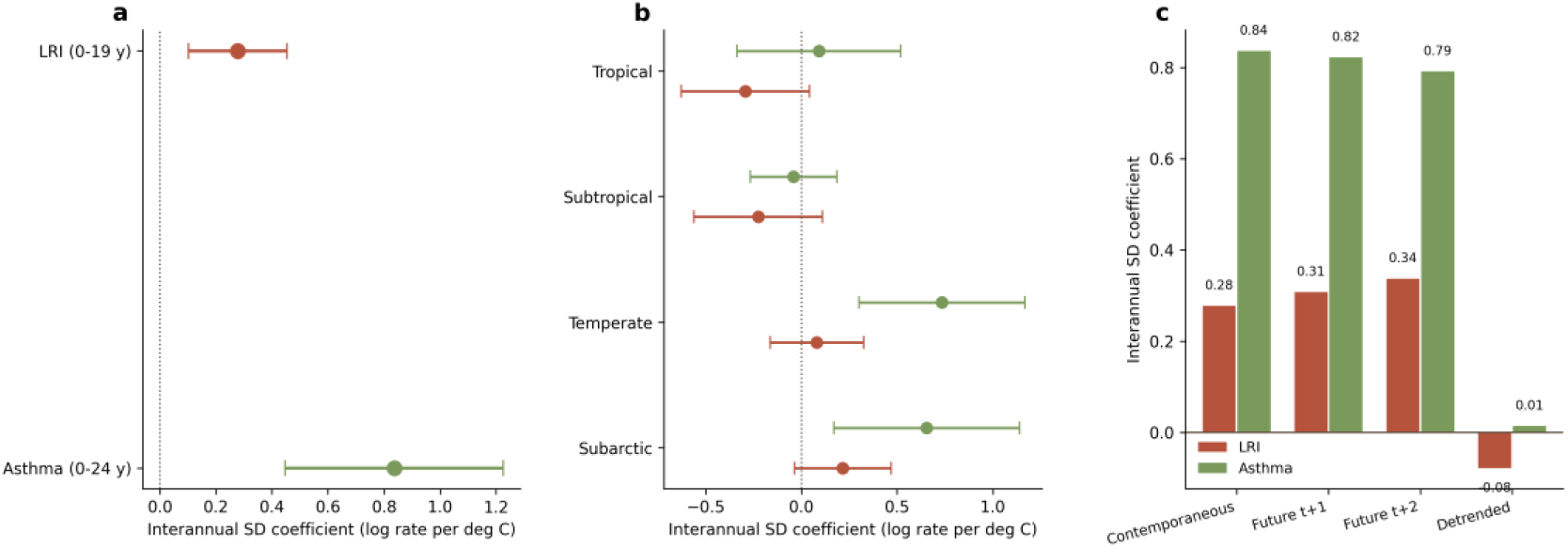
(a) Forest plot of the interannual-variability (SD) coefficient for LRI (beta 0.278, 95% CI 0.102– 0.454) and asthma (0.836, 0.447–1.226); (b) zone-specific interannual-SD coefficients for LRI and asthma (two-way FE models, Driscoll–Kraay lag 3); (c) falsification summary of the variability coefficient: contemporaneous, future-exposure (t+1, t+2) and country-detrended estimates.

**Figure 4.**
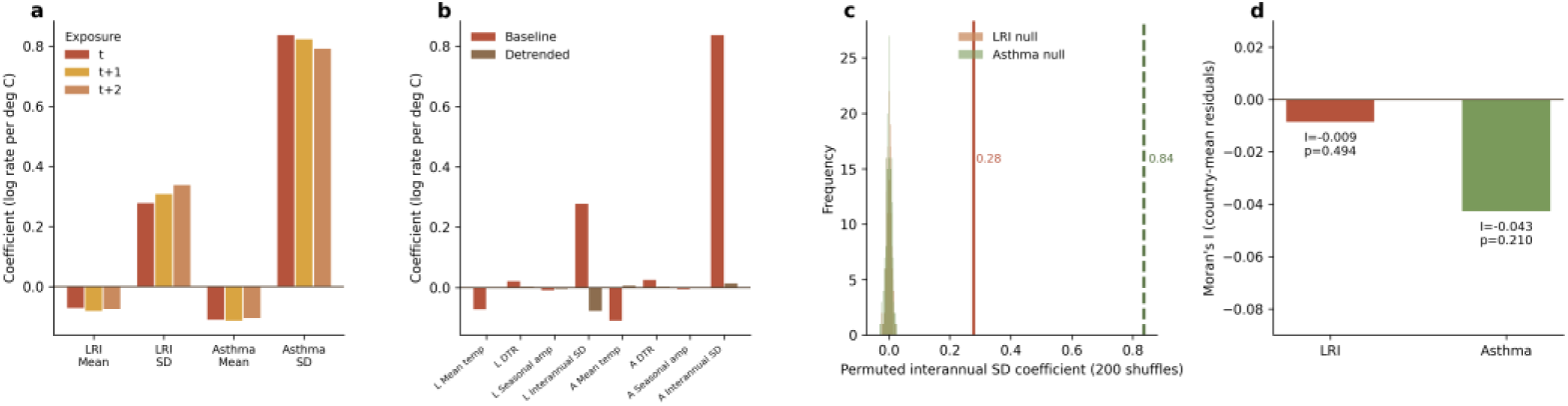
(a) Contemporaneous versus future-exposure (t+1, t+2) coefficients for mean temperature and interannual SD, both outcomes; (b) baseline versus country-linear-detrended coefficients for all four thermal metrics; (c) permutation null distributions (200 within-country year shuffles, seed 7) for the interannual-SD coefficient with observed estimates (0.28, 0.84) marked; (d) Moran’s I of country-mean residuals (LRI: I = −0.009, p = 0.494; asthma: I = −0.043, p = 0.210).

Second, within-country year-shuffle permutation (200 replicates) placed the true mean-temperature and interannual-variability coefficients outside the permuted null distribution for both outcomes (p < 0.001), confirming dependence on real temporal ordering rather than cross-sectional confounding; DTR reached marginal permutation significance (p = 0.025 for LRI; p = 0.015 for asthma) despite its null contemporaneous estimate, whereas seasonal amplitude did not (p = 0.26 and p = 0.54). Third, Moran’s I for country-mean residuals was non-significant (LRI: I = −0.009, p = 0.494; asthma: I = −0.043, p = 0.210), ruling out detectable residual spatial autocorrelation. Finally, re-estimation on the 201-country panel including the nearest-cell island states left all coefficient directions and significance unchanged. Taken together (Figure 3), the signals are temporally ordered and spatially independent but operate at the level of long-run trends, precluding contemporaneous causal inference. In the year-fixed-effect covariate model (154 countries), log GDP per capita was negatively associated (β = −0.72) and PM2.5 positively associated (β = +0.010), both statistically significant.

Adjustment for ambient PM2.5 left the main findings essentially unchanged (Figure 5)

**Figure 5.**
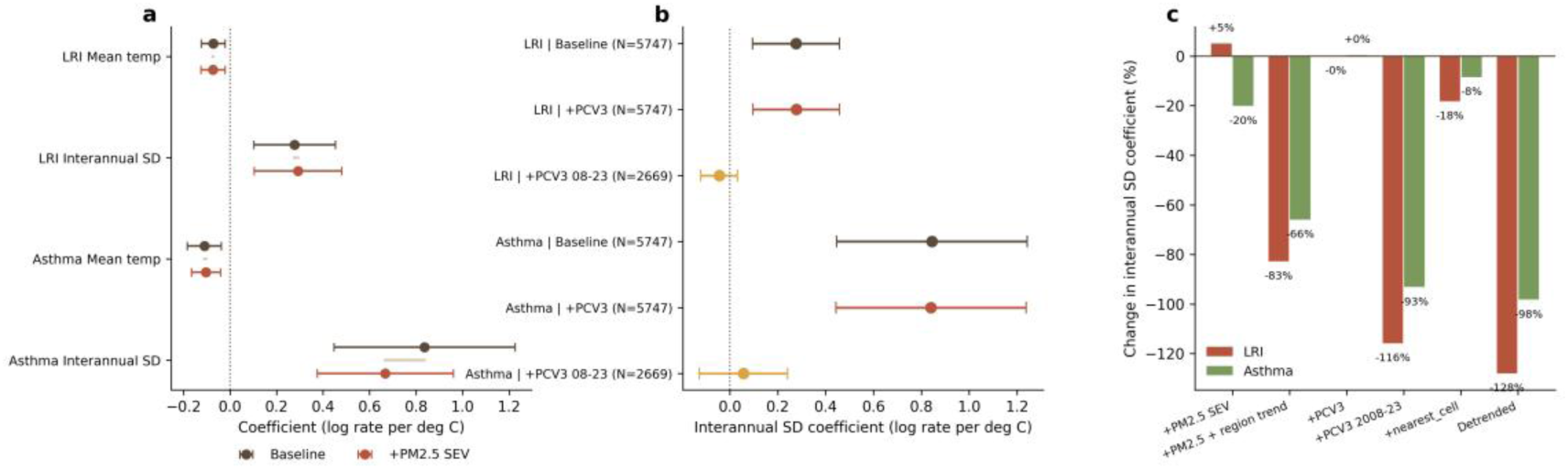
(a) Baseline versus PM2.5-SEV-adjusted coefficients for mean temperature and interannual SD, both outcomes; (b) interannual-SD coefficient under PCV3 immunization-coverage adjustment (full sample N = 5,747 and 2008–2023 subsample N = 2,669); (c) percentage change of the interannual-SD coefficient across sensitivity specifications.

Extending the falsification suite to the quadratic specification that generates the minimum mortality temperature (Supplementary Table S7) showed that the U-shape is itself trend-level. With temperatures shifted one or two years into the future, the quadratic coefficient remained positive and highly significant and the implied MMT was essentially unchanged—overall, b2 = 0.0091 (p = 1.3 × 10−11) contemporaneous versus 0.0091 (t + 1) and 0.0087 (t + 2), with MMTs of 17.1, 17.6, and 17.4 °C; tropical, b2 = 0.0181 versus 0.0173 and 0.0175 (all p < 0.001), with MMTs of 24.7, 24.6, and 25.0 °C; subtropical, b2 = 0.0128 versus 0.0117 and 0.0111 (all p ≤ 0.0003), with MMTs of 15.8, 15.4, and 14.9 °C. Future temperatures cannot generate past mortality minima, so the future-exposure check fails for the quadratic term just as it did for the linear coefficients. After country-specific linear detrending, the quadratic coefficient collapsed towards zero and lost significance overall (b2 = −0.00004; p = 0.894) and in tropical (b2 = −0.0001; p = 0.938) and temperate (b2 = −0.0003; p = 0.726) countries, and turned negative in subtropical countries (b2 = −0.0038; p = 0.040); no U-shaped form survived detrending in any zone with baseline identification. The estimated MMTs therefore locate the minimum of the long-run co-trend between warming and declining childhood mortality—a directional geographic pattern—rather than a contemporaneous optimum.

The belt-decomposition variants make the same point from the geography side (Figure 6): the U-shaped curvature persists within climate belts (model M1, country fixed effects with belt × year fixed effects: b2 = 0.0041, p = 8.8 × 10−6; implied MMT 14.3 °C), and the belt-level aggregate model (M2, belt-year means with belt and year fixed effects) gives b2 = 0.0139 (p = 0.003) with an implied MMT of 12.6 °C, whereas the between-country means model (M3: b2 = 0.0079, p = 0.24) and the belt-fixed-effects residual model (M4: b2 = −0.0014, p = 0.60) are null—consistent with the curvature living at the trend level both within and between belts.

**Figure 6.**
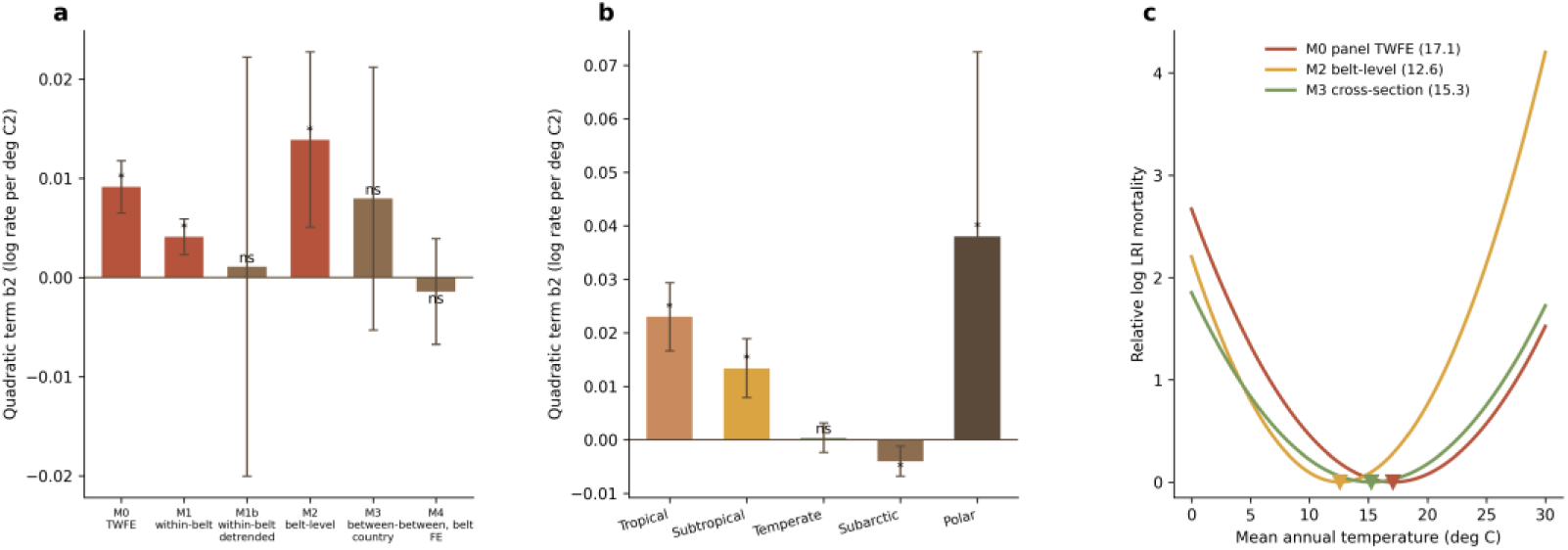
(a) Quadratic-term (b2) estimates across belt-decomposition models M0–M4 (star: p < 0.05); (b) belt × temperature interaction models (M5): b2 by climate belt; (c) implied temperature– mortality curves from the panel model (M0, MMT 17.1 °C), the belt-level model (M2, 12.6 °C) and the between-country cross-section (M3, 15.3 °C).

Adjustment for national PCV3 vaccine coverage left the main findings unchanged (Supplementary Table S8). The WUENIC PCV3 series was available for 166 of the 171 panel countries (67 country-year rows excluded; N = 5,747); years before 2008, preceding vaccine availability, were set to zero. With PCV3 coverage added to the two-way fixed-effects model, the LRI coefficients moved by less than 0.3% (interannual variability: 0.2771 to 0.2779, p = 0.0026; mean temperature: −0.0759 to −0.0757, p = 0.0057) and the asthma coefficients by less than 1% (interannual variability: 0.8441 to 0.8403, p = 3.5 × 10−5; mean temperature: −0.1154 to −0.1164, p = 0.0027); the DTR and seasonal-amplitude nulls were unaffected. PCV3 coverage itself was not associated with LRI mortality over the full period (β = 0.0001 per coverage percentage point; p = 0.58) but was inversely associated in the 2008–2023 window (β = −0.0003; p = 0.0005), consistent with its established mechanism. Restricting the adjusted model to 2008–2023 (167 countries; N = 2,669) collapsed the LRI mean-temperature and interannual-variability coefficients to null (0.0016, p = 0.80; −0.0441, p = 0.26), replicating the detrending result: the headline associations are carried by pre-2008 trend variation, and staggered pneumococcal vaccine roll-out does not account for them.

### 3.5 Spatial pattern of seasonal temperature amplitude

Country-level seasonal amplitude increases from the equator towards higher latitudes (climate-zone geography shown in Figure 1c; no country-level map is shown).

### 3.6 Projected exposure changes under CMIP6 scenarios

Across the 196 projection polygons (a climate-raster set wider than the 171-country mortality panel), matched-cell country-mean annual temperature is projected to rise by +1.61 °C under SSP1-2.6, +1.88 °C under SSP2-4.5 and +2.37 °C under SSP5-8.5 for 2041–2060 relative to 1970–2000 (Table 4). These cumulative changes span a ≈65-year interval (baseline midpoint 1985 to projection midpoint 2050) and exceed, even under the lowest scenario, the +1.1 °C warming observed over the 33-year study window 1990–2023 (Section 3.1); expressed as rates, however, only SSP5-8.5 (≈0.036 °C per year) exceeds the observed 1990–2023 rate (0.034 °C per year), whereas the SSP1-2.6 and SSP2-4.5 rates (≈0.025 and ≈0.029 °C per year) do not. Warming is greatest at high latitudes (under SSP5-8.5, Canada +4.3 °C, Russia +4.0 °C and Finland +3.4 °C), and 69% of polygons warm by more than 2 °C under SSP5-8.5. Mean diurnal temperature range changes little on average (+0.10, +0.04 and +0.04 °C under the three scenarios) and with mixed sign (39–47% of polygons show a narrowing), whereas seasonal amplitude widens in most polygons (equal-weighted means of +0.41, +0.47 and +0.43 °C; amplitude increases in 77–81% of polygons) — in contrast to the absence of an observed amplitude trend over 1990–2023. Because these are scenario-conditional climatological changes from a single climate model and the underlying mortality associations are trend-level, they quantify the direction and magnitude of future exposure change rather than any mortality response.

**Table 4.** Projected changes in country-level thermal exposure, 2041–2060 versus 1970–2000 (WorldClim v2.1 baseline and CMIP6 MRI-ESM2-0 scenarios; 196 Natural Earth admin-0 polygons, including Antarctica, dependent territories and disputed units — not identical to the 171-country mortality panel; equal-weighted polygon means)

| **Scenario** | **Δ Annual mean temperature (°C), polygon mean (5th–95th percentile)** | **Δ Mean DTR (°C), country mean** | **Polygons with narrower DTR (%)** | **Δ Seasonal amplitude (°C), country mean** | **Polygons with wider amplitude (%)** |
|---|---|---|---|---|---|
| SSP1-2.6 | +1.61 (+0.94 to +2.46) | +0.10 | 39 | +0.41 | 81 |
| SSP2-4.5 | +1.88 (+1.22 to +2.70) | +0.04 | 47 | +0.47 | 77 |
| SSP5-8.5 | +2.37 (+1.48 to +3.28) | +0.04 | 41 | +0.43 | 78 |

Note: matched-cell means over identical grid cells in the baseline and future rasters; polygons requiring buffer assignment (small island states) are excluded.

## 4. Discussion

### 4.1 Principal findings

In a 171-country panel spanning 1990–2023, this study provides the first multi-country, climate-zone-resolved estimates of the minimum mortality temperature (MMT) for childhood respiratory mortality and a decomposition of thermal variability into within-day, within-year, and between-year components. Three findings stand out. First, where identification is adequate, the childhood lower respiratory infection (LRI) MMT sits at the cool tail of the local annual-temperature distribution: 17.1 °C (95% CI 14.7 to 19.8) overall, corresponding to the 36th percentile, with 24.7 °C in tropical and 15.8 °C in subtropical countries; temperate and subarctic estimates were not meaningfully identified. Second, interannual temperature variability was positively associated with both LRI and asthma mortality, but future-exposure tests failed and country-level detrending attenuated the coefficients towards zero, so these are trend-level associations that do not support a contemporaneous causal interpretation. Third, neither mean diurnal temperature range (DTR) nor seasonal amplitude showed an independent within-country effect at the annual scale. Critically, the same falsification checks applied to the quadratic specification showed that the MMT itself is a trend-level construct: future temperatures reproduced the U-shape almost exactly, and detrending erased it in every zone with baseline identification.

### 4.2 Interpreting the child MMT

The most consequential contrast with the adult literature is positional. Adult multi-country studies place the MMT near the 60th percentile of the daily temperature distribution in tropical settings and the 80th–90th percentile in temperate ones ^1^, whereas our child estimates fall at the 21st–36th percentiles. Two non-exclusive explanations should be considered. The first is methodological: our exposure is the country-level annual mean temperature, not the daily temperature series used in adult MMT studies, so the two constructs are not directly commensurable and the percentile contrast must be read as directional rather than quantitative ^1^. The second is biological: the childhood burden of LRI mortality is concentrated in the youngest ages, and infants under one year are the most heat-vulnerable age group identified in the child-health literature ^14^. An age structure weighted towards infants would plausibly shift the mortality minimum towards cooler conditions and steepen the warm-side slope, consistent with the pattern we observe in tropical countries, where mortality rises sharply above the optimum.

Two 2025 studies bound our estimates from higher levels of the evidence hierarchy. In a Brazilian space-time-stratified case-crossover analysis of 1,061,229 under-five deaths, the daily temperature–mortality relationship was U-shaped against an explicit MMT reference, with respiratory deaths associated only with heat ^18^; our annual, cause-specific panel likewise shows the steepest warm-side slope in tropical settings, although the exposure constructs are not quantitatively commensurable. Across 56 low– and middle-income countries, monthly temperature anomalies increased under-five mortality with pronounced climate-zone heterogeneity interpreted as adaptation to local climate ^19^—the same qualitative geography our climate-zone MMTs display at the annual scale. Aggregation further separates the constructs: temporal aggregation from daily to weekly or monthly data systematically biases the tails of the exposure–response curve and the estimated MMT ^26^, and annual aggregation can only be more distortive; combined with the failed falsification of the quadratic term (Section 3.4), this is why we present the MMT geography as a directional, trend-level pattern rather than a paediatric analogue of the adult daily-scale MMT.

The policy implication is twofold. Because the child optimum lies below the typical local climate in low-latitude settings, cold-season protection for children remains important even in tropical countries, where public-health planning tends to prioritise heat; conversely, the steep warm-side slope in the tropics indicates that warming trends compound an already unfavourable thermal position. This annual-resolution evidence complements daily-scale surveillance such as the Lancet Countdown, which documents a rising number of heatwave days affecting infants ^17^: the two designs measure different exposure constructs and converge on the conclusion that young children occupy a distinct position on the temperature–mortality curve.

### 4.3 The trend-level association of interannual variability

The positive coefficients for interannual temperature variability (+0.278 for LRI; +0.836 for asthma per 1 °C) require careful framing. The future-exposure test returned nearly identical significant coefficients when mortality was regressed on temperatures one or two years ahead (0.308 at t + 1 and 0.337 at t + 2 for LRI), and country-specific detrending moved the LRI coefficient from 0.278 to −0.078. We therefore do not interpret these estimates as contemporaneous causal effects. Rather than discounting the finding as a null or spurious result, we read it as an empirical demonstration of the causal threshold of ecological annual panels: slow-moving co-trends between climate variability and declining child mortality can generate strong, temporally ordered, permutation-robust associations that nonetheless carry no same-year causal content. Confounding by co-trending air pollution does not account for the signal: controlling the national PM2.5 exposure time series shifted the interannual-variability coefficient by only +5.1% for LRI and −20.1% for asthma, with both estimates remaining significant (Section 3.4), so the trend-level association is not merely a proxy for concurrent changes in ambient air quality.

This hierarchy of evidence matters for synthesis. Daily-scale distributed lag non-linear models estimate short-term effects of temperature variability over lags of days to weeks ^13^, and multi-country applications have quantified the attendant mortality burden of non-optimal temperature ^1^, short-term variability ^7^, heatwaves ^9^, and cold spells ^10^; annual trend-level associations answer a different question and are not interchangeable with those estimates. Our results neither confirm nor exclude genuine short-term effects of interannual variability on child respiratory mortality — the present design cannot adjudicate that question, and daily-scale child-specific studies remain necessary.

### 4.4 Null findings for DTR and seasonal amplitude

The absence of independent DTR effects contrasts with adult daily time-series evidence, in which a 1 °C increase in DTR was associated with a 1.29% increase in respiratory mortality on cold days only ^4^, pooled multi-city estimates were positive (0.76% per 1 °C for respiratory mortality in the cool season) ^27^, and DTR accounted for an attributable fraction of 2.5% of mortality ^6^. The most parsimonious reconciliation is aggregation: annual mean DTR averages over the day-to-day excursions that drive acute physiological stress, diluting short-term effects below detectability in a yearly panel. The marginal permutation significance of DTR (p = 0.025 for LRI) hints at residual temporal structure but, against a null contemporaneous estimate, does not alter this interpretation. Child-specific DTR evidence remains thin and is dominated by emergency-visit outcomes rather than mortality ^15,16^; resolving whether the adult DTR signal extends to children will require daily-scale designs in this age group.

The scenario projection adds a forward-looking caveat to these annual-scale nulls: seasonal amplitude is projected to widen in roughly four out of five polygons under all three SSP scenarios (Section 3.6), so the thermal-variability regime of the coming decades may differ from the 1990– 2023 window analysed here, even where no within-country association was detectable historically.

### 4.5 Limitations

including baseline development and geography that mediate long-run climate–health relationships (the time-varying pollution and immunization covariates were adjusted separately in §3.4 and §3.6); our estimates are therefore identified^26^

## 5. Conclusions

Across 171 countries over 1990–2023, this study delivers two contributions. First, it provides the first multi-country, climate-zone-resolved geography of the childhood respiratory minimum mortality temperature under an annual construct: 17.1 °C (95% CI 14.7–19.8) overall—at the 36th percentile of the annual-temperature distribution—with 24.7 °C in tropical and 15.8 °C in subtropical countries, placing the childhood optimum at the cool tail of local climates; this is a directional, trend-level pattern, as the underlying quadratic association did not survive detrending. Second, interannual temperature variability was positively associated with childhood lower respiratory infection (+0.278 per 1 °C) and asthma mortality (+0.836 per 1 °C), yet failed future-exposure falsification restricts these to trend-level associations; neither diurnal temperature range nor seasonal amplitude showed independent annual effects. The practical takeaway is that children occupy a distinct, cooler position on the temperature–mortality curve than adults (directional only, given differing temperature constructs) ^1^, so at the trend level, cold-season protection remains a relevant consideration even in the tropics. Future work should apply daily-scale DTR designs to paediatric outcomes, extend analyses to subnational and country-specific mortality series, and couple the estimated response functions with CMIP6 climate scenarios to project child respiratory burden under warming. A first step is included in this version: the matched-cell CMIP6 projection of Sections 2.8 and 3.6 quantifies country-level exposure changes to 2041–2060 (country-mean warming of +1.6 to +2.4 °C, with seasonal amplitude widening in most polygons) without extrapolating the trend-level coefficients to mortality; extension to multi-model ensembles is the appropriate next step.

## Declarations

### Ethics approval and consent to participate

Not applicable. The study used publicly available, de-identified, aggregated estimates and was exempt from ethics review.

### Consent for publication

Not applicable.

### Availability of data and materials

Mortality estimates are publicly available from the Global Burden of Disease Results Tool (https://vizhub.healthdata.org/gbd-results) ^24^. The C-LSAT HRv1 and C-LDTR HRv1 climate datasets are available on figshare (https://doi.org/10.6084/m9.figshare.28255505 and https://doi.org/10.6084/m9.figshare.28255568) ^20^. The analysis code and the analytical panel are available from the corresponding author upon reasonable request. The WorldClim v2.1 baseline and CMIP6-downscaled scenario fields are publicly available (https://www.worldclim.org; https://geodata.ucdavis.edu/cmip6); the per-country projection table (CSV) is provided as Additional file 2; the raster-extraction script used to produce it is available from the corresponding author upon reasonable request.

Funding This work was supported by the Beijing Science and Technology Nova Program Interdisciplinary Project (20230484439) and the Beijing High-Level Public Health Technical Talents Project (2023-02-33). The funder had no role in study design, data collection, data analysis, data interpretation, or writing of the report.

### Presentation

This work has not been presented at any scientific meeting.

Disclosure The authors declare no conflicts of interest. AI tools were used for language editing and for assistance with code generation and modification; the authors take full responsibility for the content.

Authors’ contributions SC conceived the study, led the research design, and is the principal investigator responsible for the study idea and overall conduct. DL and HC curated the data and performed the formal analysis. DL drafted the manuscript. SC supervised the study and is the corresponding author. All authors read and approved the final manuscript.

## Data Availability

Availability of data and materials Mortality estimates are publicly available from the Global Burden of Disease Results Tool (https://vizhub.healthdata.org/gbd-results) 24. The C-LSAT HRv1 and C-LDTR HRv1 climate datasets are available on figshare (https://doi.org/10.6084/m9.figshare.28255505 and https://doi.org/10.6084/m9.figshare.28255568) 20. The analysis code and the analytical panel are available from the corresponding author upon reasonable request. The WorldClim v2.1 baseline and CMIP6-downscaled scenario fields are publicly available (https://www.worldclim.org; https://geodata.ucdavis.edu/cmip6); the per-country projection table (CSV) is provided as Additional file 3; the raster-extraction script used to produce it is available from the corresponding author upon reasonable request.

https://cds.climate.copernicus.eu/

## Acknowledgements

We thank the Institute for Health Metrics and Evaluation and the Global Burden of Disease collaborative network for making cause-specific mortality estimates publicly available. We also thank the C-LSAT team at Sun Yat-sen University for developing and sharing the high-resolution land surface temperature and diurnal temperature range datasets.

## Additional files

**Additional file 1: Supplementary Tables S1–S8.** Panel description (S1); main model coefficients (S2); MMT estimates by climate zone with bootstrap details (S3); falsification suite results (S4); descriptive trends of thermal metrics (S5); PM2.5-adjustment comparison (S6); falsification of the quadratic MMT specification (S7); WUENIC PCV3-adjustment comparison (S8).

Additional file 2: Country-level SSP scenario exposure projections (196 Natural Earth admin-0 polygons × 3 scenarios, including Antarctica, dependent territories and disputed units — not identical to the 171-country mortality panel; matched-cell changes in annual mean temperature, diurnal temperature range and seasonal amplitude).

## Notes

### Competing Interest Statement

The authors have declared no competing interest.

### Summary of Updates

An internal validation against LRI death counts was added; the Figure 4 description was rewritten; limitations expanded. The reporting-checklist (GATHER) sentence and checklist file were removed with references renumbered; 'Fig.' abbreviations expanded and glued citations separated. Funding and AI-use declarations updated.

