## Supplementary_Materials for "Thermal variability and the geography of optimal temperature for child survival: childhood respiratory-infection mortality in 171 countries: a systematic analysis of the Global Burden of Disease Study 2023 and the C-LSAT high-resolution climate dataset"

*Analytical scope: Global Burden of Disease Study 2023 (GBD 2023, v8352) × C-LSAT HRv1 high-resolution climate data; lower respiratory infection (LRI) deaths at ages 0–19 years and asthma deaths at ages 0–24 years; 171 countries, 1990–2023; outcome measure: deaths; two-way fixed-effects (TWFE) models.*

Contents: Supplementary Table S1 (panel description); Supplementary Table S2 (main two-way fixed-effects coefficients); Supplementary Table S3 (minimum mortality temperature by climate zone); Supplementary Table S4 (falsification and sensitivity suite); Supplementary Table S5 (descriptive trends of thermal metrics, 1990–2023); Supplementary Table S6 (robustness to adjustment for ambient PM2.5 exposure); Supplementary Table S7 (falsification of the quadratic minimum-mortality-temperature specification); Supplementary Table S8 (robustness to adjustment for WUENIC PCV3 vaccine coverage); Supplementary Table S9 (between/within climate-zone decomposition of the quadratic U-shape).

#### Supplementary Table S1. Description of the analytical panel: country × year structure, composition of the 171-country main panel, and missing-data structure.

| Item | LRI (ages 0–19 years) | Asthma (ages 0–24 years) |
|---|---|---|
| GBD 2023 countries/territories | 204 | 204 |
| Countries matched to climate data | 201 | 201 |
| of which polygon-masked (main panel) | 171 | 171 |
| of which nearest_cell small-island states (sensitivity analysis only) | 30 | 30 |
| GBD countries unmatched to the climate file | France, Norway, Tokelau | France, Norway, Tokelau |
| Country-year observations (main panel) | 5,814 | 5,814 |
| Study period | 1990–2023 | 1990–2023 |
| Missing fixed 2022 population denominator (main panel) | Greenland; Saint Vincent and the Grenadines; Taiwan; United States Virgin Islands (4 countries) | Greenland; Saint Vincent and the Grenadines; Taiwan; United States Virgin Islands (4 countries) |
| Missing covariates (log GDP per capita, PM2.5, urban population share) | 17 countries | 17 countries |

*Note. The main panel is balanced: 171 countries × 34 years (1990–2023) = 5,814 country-year observations per outcome.*

*The 171-country set is the structural maximum of the GBD ∩ polygon-climate intersection: six polygon-covered climate territories (Cayman Islands, Falkland Islands, New Caledonia, South Georgia, Western Sahara, Åland) have no GBD estimates, and three GBD countries (France, Norway, Tokelau) have no records in the climate file.*

*Mortality rates use the GLOBOCAN 2022 population aged 0–19 years as a fixed denominator per 100,000 for all years. For the four countries lacking this denominator, the time-invariant constant is absorbed by country fixed effects, so fixed-effect coefficients are unaffected; these countries are excluded from rate-level descriptive statistics.*

*The covariates (log GDP per capita, PM2.5, urban population share) are time-varying; the year-fixed-effect covariate model (154 countries; no country fixed effects, so coefficients reflect both cross-sectional and temporal variation) includes them in addition to the four thermal exposures (Supplementary Table S4).*

**Supplementary Table S2. Main two-way fixed-effects models: coefficients (95% CI) and p-values for four thermal exposures and two childhood mortality outcomes, 171 countries, 1990–2023.**

| Exposure | LRI 0–19: coefficient (95% CI) | LRI 0–19: p | Asthma 0–24: coefficient (95% CI) | Asthma 0–24: p |
|---|---|---|---|---|
| Annual mean temperature (°C) | −0.072 (−0.124 – −0.020) | 0.006 | −0.110 (−0.183 – −0.037) | 0.003 |
| Diurnal temperature range (°C) | 0.020 (−0.047 – 0.088) | 0.558 | 0.025 (−0.071 – 0.121) | 0.612 |
| Seasonal amplitude (°C) | −0.009 (−0.021 – 0.004) | 0.194 | −0.006 (−0.025 – 0.014) | 0.567 |
| Interannual variability (10-year SD of annual mean, °C) | 0.278 (0.102 – 0.454) | 0.002 | 0.836 (0.447 – 1.226) | $2.6\times10^{-5}$ |
| Within $R^2$ | 0.097 | — | 0.166 | — |
| Country-years (N) / countries | 5,814 / 171 | — | 5,814 / 171 | — |

*Note. Model: log(mortality rate) ~ mean_temp + dtr_mean + season_amp + interannual_sd with country and year fixed effects; Driscoll–Kraay standard errors, bandwidth lag = 3. Coefficients are log-rate changes per 1 °C; exponentiated, a 1 °C increase in interannual variability corresponds to +32.0% (LRI) and +130.8% (asthma) mortality. p-values <0.001 are reported exactly. The falsification suite (Supplementary Table S4) shows these associations are trend-level co-movements and do not support a contemporaneous causal interpretation.*

**Supplementary Table S3. Minimum mortality temperature (MMT) by climate zone, with bootstrap confidence intervals, position in the zone temperature distribution, and number of valid bootstrap replications.**

| Climate zone | Countries (n) | Country-years (N) | MMT (°C) | 95% CI (°C) | MMT percentile of zone temperature distribution | Valid bootstrap replications (of 300) |
|---|---|---|---|---|---|---|
| Overall | 171 | 5,814 | 17.1 | 14.7 – 19.8 | 36th | 300 |
| Tropical | 86 | 2,924 | 24.7 | 18.5 – 27.0 | 32nd | 289 |
| Subtropical | 29 | 986 | 15.8 | 3.1 – 20.1 | 21st | 285 |
| Temperate | 46 | 1,564 | 25.5 | 8.1 – 28.3 | 100th | 181 |
| Subarctic | 9 | 306 | Not identified | — | — | 84 |

*Note. MMT is the turning point −b1/(2 × b2) of a quadratic term in annual mean temperature added to the main model; b2 > 0 indicates a U-shaped association. Confidence intervals are percentile intervals from 300 country-cluster bootstrap replications; "valid bootstrap replications" counts replicates yielding an identified U-shaped minimum.*

*Temperate zone: the point estimate falls at the 100th percentile of the zone temperature distribution (upper bound) and only 181 of 300 bootstrap replicates were valid; identification is weak and the estimate should be read with caution.*

*Subarctic zone: the quadratic coefficient is negative (inverted-U), so no mortality minimum exists; the MMT is not identified. Across the 84 valid bootstrap replicates the turning points ranged from 2.6 to 38.7 °C, indicating instability.*

*Climate-zone rows sum to 170 countries: Greenland is not assigned to any climate zone in the source classification and therefore contributes to the overall estimate only.*

*The exposure is the country-level annual mean temperature, not the daily temperature distributions used in adult MMT studies; percentile comparisons with adult estimates are directional only.*

**Supplementary Table S4. Falsification and sensitivity suite: future-exposure leads, country-level detrending, permutation tests, Moran's I, the 201-country sensitivity panel, and the 154-country covariate model.**

| Check | Term | LRI (ages 0–19) | Asthma (ages 0–24) | N |
|---|---|---|---|---|
| Future exposure t+1 | Annual mean temperature | β = −0.081 (p = 0.006) | β = −0.114 (p = 0.003) | 5,643 |
| | Diurnal temperature range | β = 0.016 (p = 0.657) | β = 0.013 (p = 0.783) | |
| | Seasonal amplitude | β = −0.006 (p = 0.324) | β = −0.002 (p = 0.856) | |
| | Interannual variability | β = 0.308 (p < 0.001) | β = 0.822 (p < 0.001) | |
| Future exposure t+2 | Annual mean temperature | β = −0.075 (p = 0.009) | β = −0.105 (p = 0.008) | 5,472 |
| | Diurnal temperature range | β = 0.010 (p = 0.782) | β = −0.002 (p = 0.961) | |
| | Seasonal amplitude | β = −0.007 (p = 0.304) | β = −0.001 (p = 0.944) | |
| | Interannual variability | β = 0.337 (p < 0.001) | β = 0.791 (p < 0.001) | |
| Country linear detrending | Annual mean temperature | β = −0.002 (p = 0.806) | β = 0.006 (p = 0.335) | 5,814 |
| | Diurnal temperature range | β = 0.004 (p = 0.597) | β = 0.003 (p = 0.536) | |
| | Seasonal amplitude | β = −0.004 (p = 0.045) | β = 0.0002 (p = 0.923) | |
| | Interannual variability | β = −0.078 (p = 0.214) | β = 0.014 (p = 0.649) | |
| Permutation test (200 within-country year shuffles) | Annual mean temperature | β = −0.072; p < 0.005 | β = −0.110; p < 0.005 | 200 |
| | Diurnal temperature range | β = 0.020; p = 0.025 | β = 0.025; p = 0.015 | |
| | Seasonal amplitude | β = −0.009; p = 0.260 | β = −0.006; p = 0.535 | |
| | Interannual variability | β = 0.278; p < 0.005 | β = 0.836; p < 0.005 | |
| Moran's I (KNN k = 5, country-mean residuals) | I statistic | I = −0.009 (p = 0.494) | I = −0.043 (p = 0.210) | 167 |
| 201-country sensitivity (adds 30 nearest_cell states) | Annual mean temperature | β = −0.064 (p = 0.007) | β = −0.099 (p = 0.004) | 6,834 |
| | Diurnal temperature range | β = 0.018 (p = 0.560) | β = 0.027 (p = 0.529) | |
| | Seasonal amplitude | β = −0.008 (p = 0.186) | β = −0.006 (p = 0.551) | |
| | Interannual variability | β = 0.227 (p = 0.008) | β = 0.765 (p < 0.001) | |
| 154-country covariate model (year FE, no country FE) | Annual mean temperature | β = −0.026 (p < 0.001) | β = −0.021 (p < 0.001) | 5,236 |

| | | |
|---|---|---|
| Diurnal temperature range | β = −0.028 (p < 0.001) | β = −0.0002 (p = 0.977) |
| Seasonal amplitude | β = −0.036 (p < 0.001) | β = −0.094 (p < 0.001) |
| Interannual variability | β = −0.108 (p = 0.757) | β = −0.100 (p = 0.707) |
| log GDP per capita | β = −0.724 (p < 0.001) | β = −0.186 (p < 0.001) |
| PM2.5 | β = 0.010 (p < 0.001) | β = −0.0004 (p = 0.682) |
| Urban population share | β = −0.004 (p < 0.001) | β = −0.001 (p = 0.015) |

*Note. β is the log-rate coefficient per 1 °C. Permutation p-values are the fraction of shuffled statistics exceeding the true estimate in absolute value (resolution 1/200 = 0.005).*

*Future-exposure (t+1, t+2) models return coefficients nearly identical to, and as significant as, the contemporaneous model: future temperature "predicts" current mortality, so the falsification is not passed and a contemporaneous causal interpretation is not supported.*

*After country-specific linear detrending, all coefficients attenuate toward zero and lose significance, consistent with trend-level co-movement. Permutation tests show the associations depend on the true temporal ordering (except seasonal amplitude), jointly supporting a trend-level rather than a year-to-year interpretation.*

*Moran's I of country-mean residuals is not significant for either outcome; a spatial error model is not required. Re-including the 30 nearest_cell small-island states (201 countries) leaves signs and significance unchanged. The covariate model uses year fixed effects only (no country fixed effects), so its coefficients reflect both cross-sectional and temporal variation (154 of 171 countries have complete covariates; N = 5,236).*

### Supplementary Table S5. Descriptive trends of the four thermal metrics, 1990–2023 (equal-weighted country means, 171 countries), and 2023 cross-sectional correlations.

| Metric | Slope (°C per year) | SE | p | 1990 value (°C) | 2023 value (°C) |
|---|---|---|---|---|---|
| Annual mean temperature | 0.0337 | 0.0025 | $1.4\times10^{-14}$ | 18.83 | 19.92 |
| Diurnal temperature range | 0.0084 | 0.0011 | $5.4\times10^{-9}$ | 10.32 | 10.45 |
| Seasonal amplitude | 0.0024 | 0.0060 | 0.691 | 12.53 | 12.71 |
| Interannual variability (10-year SD of annual mean, °C) | −0.0024 | 0.0005 | $8.5\times10^{-5}$ | 0.42 | 0.35 |
| **2023 cross-section (n = 167 countries): Spearman ρ of the log LRI mortality rate against each metric** | | | | | |
| Seasonal amplitude | ρ = −0.424 | — | $1.2\times10^{-8}$ | — | — |
| Diurnal temperature range | ρ = 0.443 | — | $2.0\times10^{-9}$ | — | — |

*Note. Slopes are ordinary least-squares trends of the equal-weighted 171-country annual means on calendar year; endpoint values are the 1990 and 2023 annual means. Countries became warmer on average (+0.0337 °C/year), with marginally wider day–night contrast, slightly damped year-to-year variability, and a stable seasonal cycle.*

*The cross-sectional correlations (log LRI mortality rate vs seasonal amplitude ρ = −0.424; vs diurnal temperature range ρ = +0.443) describe between-country variation in 2023 and differ in sign from the within-country fixed-effects estimates; the two are not contradictory because the fixed-effects model identifies within-country variation only.*

### Supplementary Table S6. Robustness to adjustment for ambient PM2.5 exposure (GBD 2023 summary exposure value, SEV, country–year series): baseline versus PM2.5-adjusted two-way fixed-effect coefficients.

| Outcome | Model | Variable | Coef. | 95% CI | p | Within $R^2$ | Δ vs baseline |
|---|---|---|---|---|---|---|---|

| | | | | | | | (%) |
|---|---|---|---|---|---|---|---|
| LRI (0–19) | Baseline (TWFE) | Annual mean temperature | −0.0721 | (−0.1239 – −0.0203) | 0.0063 | 0.097 | — |
| LRI (0–19) | Baseline (TWFE) | Diurnal temperature range | 0.0202 | (−0.0474 – 0.0878) | 0.558 | 0.097 | — |
| LRI (0–19) | Baseline (TWFE) | Seasonal amplitude | −0.0085 | (−0.0214 – 0.0043) | 0.194 | 0.097 | — |
| LRI (0–19) | Baseline (TWFE) | Interannual SD | 0.2780 | (0.1019 – 0.4540) | 0.0020 | 0.097 | — |
| LRI (0–19) | +PM2.5 SEV | Annual mean temperature | −0.0727 | (−0.1242 – −0.0212) | 0.0056 | 0.100 | −0.8 |
| LRI (0–19) | +PM2.5 SEV | Diurnal temperature range | 0.0185 | (−0.0520 – 0.0889) | 0.607 | 0.100 | −8.7 |
| LRI (0–19) | +PM2.5 SEV | Seasonal amplitude | −0.0084 | (−0.0216 – 0.0048) | 0.214 | 0.100 | +2.0 |
| LRI (0–19) | +PM2.5 SEV | Interannual SD | 0.2923 | (0.1042 – 0.4803) | 0.0023 | 0.100 | +5.1 |
| LRI (0–19) | +PM2.5 SEV | PM2.5 SEV | −0.0018 | (−0.0043 – 0.0008) | 0.181 | 0.100 | — |
| LRI (0–19) | +PM2.5 SEV + region × year trend | Annual mean temperature | −0.0114 | (−0.0312 – 0.0084) | 0.258 | −0.896 | +84.2 |
| LRI (0–19) | +PM2.5 SEV + region × year trend | Diurnal temperature range | 0.0018 | (−0.0253 – 0.0288) | 0.899 | −0.896 | −91.3 |
| LRI (0–19) | +PM2.5 SEV + region × year trend | Seasonal amplitude | −0.0025 | (−0.0063 – 0.0014) | 0.213 | −0.896 | +71.2 |
| LRI (0–19) | +PM2.5 SEV + region × year trend | Interannual SD | 0.0478 | (−0.0817 – 0.1774) | 0.469 | −0.896 | −82.8 |
| LRI (0–19) | +PM2.5 SEV + region × year trend | PM2.5 SEV | −0.0116 | (−0.0136 – −0.0097) | <0.001 | −0.896 | — |
| Asthma (0–24) | Baseline (TWFE) | Annual mean temperature | −0.1102 | (−0.1829 – −0.0374) | 0.0030 | 0.166 | — |
| Asthma (0–24) | Baseline (TWFE) | Diurnal temperature range | 0.0249 | (−0.0712 – 0.1210) | 0.612 | 0.166 | — |
| Asthma (0–24) | Baseline (TWFE) | Seasonal amplitude | −0.0058 | (−0.0255 – 0.0140) | 0.567 | 0.166 | — |
| Asthma (0–24) | Baseline (TWFE) | Interannual SD | 0.8363 | (0.4468 – 1.2258) | $2.6\times10^{-5}$ | 0.166 | — |
| Asthma (0–24) | +PM2.5 SEV | Annual mean temperature | −0.1031 | (−0.1659 – −0.0402) | 0.0013 | 0.182 | +6.4 |

| | | | | | | | |
|---|---|---|---|---|---|---|---|
| Asthma (0–24) | +PM2.5 SEV | Diurnal temperature range | 0.0456 | (−0.0420 – 0.1333) | 0.308 | 0.182 | +83.2 |
| Asthma (0–24) | +PM2.5 SEV | Seasonal amplitude | −0.0078 | (−0.0253 – 0.0098) | 0.385 | 0.182 | −34.9 |
| Asthma (0–24) | +PM2.5 SEV | Interannual SD | 0.6679 | (0.3755 – 0.9604) | $7.7\times10^{-6}$ | 0.182 | −20.1 |
| Asthma (0–24) | +PM2.5 SEV | PM2.5 SEV | 0.0208 | (0.0176 – 0.0240) | <0.001 | 0.182 | — |
| Asthma (0–24) | +PM2.5 SEV + region × year trend | Annual mean temperature | −0.0163 | (−0.0538 – 0.0213) | 0.396 | −1.094 | +85.2 |
| Asthma (0–24) | +PM2.5 SEV + region × year trend | Diurnal temperature range | 0.0146 | (−0.0180 – 0.0472) | 0.379 | −1.094 | −41.3 |
| Asthma (0–24) | +PM2.5 SEV + region × year trend | Seasonal amplitude | 0.0007 | (−0.0061 – 0.0075) | 0.841 | −1.094 | +112.1 |
| Asthma (0–24) | +PM2.5 SEV + region × year trend | Interannual SD | 0.2844 | (0.1088 – 0.4600) | 0.0015 | −1.094 | −66.0 |
| Asthma (0–24) | +PM2.5 SEV + region × year trend | PM2.5 SEV | 0.0075 | (0.0045 – 0.0105) | $7.9\times10^{-7}$ | −1.094 | — |

*Note. SEV is a modelled 0–100 summary exposure value for ambient particulate matter pollution (GBD 2023); the platform does not provide µg/m³ concentrations. The third model adds GBD super-region × year trends. Coefficients are log-rate effects per 1 °C (thermal metrics) or per SEV unit (PM2.5 SEV). Δ vs baseline is the percentage change in the coefficient relative to the baseline model. All models use country and year fixed effects with Driscoll–Kraay standard errors (bandwidth lag 3); N = 5,814 country-years (171 countries).*

### Supplementary Table S7. Falsification of the quadratic specification underlying the minimum mortality temperature (MMT): quadratic coefficient b2 and implied MMT under contemporaneous, future-exposure (t+1, t+2), and country-linear-detrended models, overall and by climate zone.

| Climate zone | Model | b1 (linear term) | b2 (quadratic term, 95% CI) | b2 p | MMT (°C) | N |
|---|---|---|---|---|---|---|
| Overall | Contemporaneous | -0.3122 | 0.00913 (0.00649 – 0.01177) | $1.3\times10^{-11}$ | 17.1 | 5,814 |
| | Future exposure t+1 | -0.3202 | 0.00911 (0.00647 – 0.01175) | $1.4\times10^{-11}$ | 17.6 | 5,643 |
| | Future exposure t+2 | -0.3048 | 0.00873 (0.00614 – 0.01133) | $4.3\times10^{-11}$ | 17.4 | 5,472 |
| | Country-linear detrended | -0.0011 | −0.00004 (−0.00057 – 0.00050) | 0.894 | Not identified | 5,814 |
| Tropical | Contemporaneous | -0.8931 | 0.01811 (0.01384 – 0.02239) | $2.2\times10^{-16}$ | 24.7 | 2,924 |
| | Future exposure t+1 | -0.8523 | 0.01730 (0.01251 – 0.02208) | $1.7\times10^{-12}$ | 24.6 | 2,838 |
| | Future exposure t+2 | -0.8737 | 0.01750 (0.01200 – 0.02299) | $4.8\times10^{-10}$ | 25.0 | 2,752 |

| | | | | | | |
|---|---|---|---|---|---|---|
| | Country-linear detrended | 0.0008 | −0.00013 (−0.00338 – 0.00312) | 0.938 | Not identified | 2,924 |
| Subtropical | Contemporaneous | -0.4017 | 0.01275 (0.00783 – 0.01767) | $4.4\times10^{-7}$ | 15.8 | 986 |
| | Future exposure t+1 | -0.3600 | 0.01170 (0.00640 – 0.01700) | $1.6\times10^{-5}$ | 15.4 | 957 |
| | Future exposure t+2 | -0.3330 | 0.01114 (0.00519 – 0.01709) | $2.5\times10^{-4}$ | 14.9 | 928 |
| | Country-linear detrended | 0.1561 | −0.00380 (−0.00743 – −0.00017) | 0.040 | Not identified | 986 |
| Temperate | Contemporaneous | -0.0715 | 0.00140 (−0.00102 – 0.00383) | 0.256 | 25.5 | 1,564 |
| | Future exposure t+1 | -0.0763 | 0.00160 (−0.00064 – 0.00384) | 0.161 | 23.8 | 1,518 |
| | Future exposure t+2 | -0.0851 | 0.00226 (−0.00014 – 0.00467) | 0.065 | 18.8 | 1,472 |
| | Country-linear detrended | 0.0030 | −0.00034 (−0.00222 – 0.00155) | 0.726 | Not identified | 1,564 |
| Subarctic | Contemporaneous | -0.0349 | −0.00297 (−0.00689 – 0.00095) | 0.137 | Not identified | 306 |
| | Future exposure t+1 | -0.0398 | −0.00407 (−0.00753 – −0.00061) | 0.021 | Not identified | 297 |
| | Future exposure t+2 | -0.0422 | −0.00424 (−0.00732 – −0.00115) | 0.007 | Not identified | 288 |
| | Country-linear detrended | -0.0069 | 0.00055 (−0.00171 – 0.00282) | 0.630 | 6.2 | 306 |

*Note. Model: log(mortality rate) ~ mean_temp + mean_temp² + dtr_mean + season_amp + interannual_sd with country and year fixed effects (Driscoll–Kraay standard errors, bandwidth 3); the detrended model applies country-specific linear detrending to all variables (including mean_temp²) with year fixed effects only. MMT = −b1/(2 × b2), shown only where b2 > 0.*

*Future-exposure models (t+1, t+2) return essentially unchanged, highly significant quadratic coefficients and MMTs (e.g., overall MMT 17.1, 17.6, 17.4 °C), so the future-exposure falsification fails for the quadratic term: the U-shape is carried by slow-moving co-trends. After country-linear detrending, b2 collapses to zero and loses significance overall (p = 0.894) and in the tropical (p = 0.938) and temperate (p = 0.726) zones, and turns negative in the subtropical zone (p = 0.040); no U-shaped form survives detrending in any zone with baseline identification. The MMT estimates in Table 3 and Supplementary Table S3 are therefore trend-level, directional patterns of the annual construct.*

### Supplementary Table S8. Robustness to adjustment for national PCV3 vaccine coverage (WHO/UNICEF estimates of national immunization coverage, WUENIC 2025 revision): baseline versus PCV3-adjusted two-way fixed-effect coefficients, 1990–2023 (pre-2008 coverage set to zero) and restricted to 2008–2023.

| Outcome | Model | Variable | Coef. | 95% CI | p | Δ vs baseline (%) | N |
|---|---|---|---|---|---|---|---|
| LRI (0–19) | Baseline (TWFE) | Annual mean temperature | −0.0759 | (−0.1296 – −0.0223) | 0.006 | — | 5,747 |
| | | Diurnal temperature range | 0.0206 | (−0.0452 – 0.0863) | 0.540 | — | |
| | | Seasonal amplitude | −0.0088 | (−0.0220 – 0.0044) | 0.192 | — | |
| | | Interannual | 0.2771 | (0.0957 – 0.4584) | 0.003 | — | |

| | | | | | | | |
|---|---|---|---|---|---|---|---|
| | | variability | | | | | |
| | +PCV3 coverage | Annual mean temperature | −0.0757 | (−0.1293 – −0.0221) | 0.006 | +0.3 | 5,747 |
| | | Diurnal temperature range | 0.0206 | (−0.0453 – 0.0864) | 0.540 | +0.1 | |
| | | Seasonal amplitude | −0.0088 | (−0.0219 – 0.0044) | 0.192 | +0.0 | |
| | | Interannual variability | 0.2779 | (0.0974 – 0.4584) | 0.003 | +0.3 | |
| | | PCV3 coverage (%) | 0.0001 | (−0.0003 – 0.0006) | 0.581 | — | |
| | +PCV3 coverage, 2008–2023 sample | Annual mean temperature | 0.0016 | (−0.0108 – 0.0140) | 0.798 | +102.1 | 2,669 |
| | | Diurnal temperature range | −0.0407 | (−0.0726 – −0.0088) | 0.012 | −297.9 | |
| | | Seasonal amplitude | 0.0043 | (−0.0016 – 0.0101) | 0.151 | +148.5 | |
| | | Interannual variability | −0.0441 | (−0.1205 – 0.0323) | 0.257 | −115.9 | |
| | | PCV3 coverage (%) | −0.0003 | (−0.0005 – −0.0001) | $4.8\times10^{-4}$ | — | |
| Asthma (0–24) | Baseline (TWFE) | Annual mean temperature | −0.1154 | (−0.1916 – −0.0393) | 0.003 | — | 5,747 |
| | | Diurnal temperature range | 0.0221 | (−0.0702 – 0.1143) | 0.639 | — | |
| | | Seasonal amplitude | −0.0058 | (−0.0259 – 0.0143) | 0.573 | — | |
| | | Interannual variability | 0.8441 | (0.4466 – 1.2416) | $3.2\times10^{-5}$ | — | |
| | +PCV3 coverage | Annual mean temperature | −0.1164 | (−0.1923 – −0.0405) | 0.003 | −0.8 | 5,747 |
| | | Diurnal temperature range | 0.0220 | (−0.0699 – 0.1139) | 0.639 | −0.3 | |
| | | Seasonal amplitude | −0.0058 | (−0.0260 – 0.0144) | 0.575 | −0.1 | |
| | | Interannual variability | 0.8403 | (0.4428 – 1.2378) | $3.5\times10^{-5}$ | −0.5 | |
| | | PCV3 coverage (%) | −0.0006 | (−0.0011 – −0.0000) | 0.034 | — | |
| | +PCV3 coverage, 2008–2023 sample | Annual mean temperature | −0.0275 | (−0.0452 – −0.0098) | 0.002 | +76.1 | 2,669 |
| | | Diurnal temperature range | −0.0728 | (−0.1061 – −0.0394) | $1.9\times10^{-5}$ | −429.6 | |
| | | Seasonal amplitude | 0.0040 | (−0.0125 – 0.0206) | 0.633 | +169.8 | |
| | | Interannual variability | 0.0571 | (−0.1273 – 0.2415) | 0.544 | −93.2 | |
| | | PCV3 coverage (%) | 0.0003 | (−0.0001 – 0.0007) | 0.098 | — | |

*Note. TWFE = two-way fixed effects (country + year), Driscoll–Kraay standard errors, bandwidth 3. PCV3 = third-dose pneumococcal conjugate vaccine coverage (%), WUENIC 2025 revision (WHO Global Health Observatory, retrieved 28 August 2026); the series starts in 2008, so 1990–2007 values (pre-availability) are set to zero. Five panel countries lack a WUENIC PCV3 series (Greenland, Puerto Rico, South Sudan, Taiwan, United States Virgin Islands; 67 country-year rows excluded), giving N = 5,747; the 2008–2023 restricted sample has 167 countries and N = 2,669.*

*Adding PCV3 coverage changes every temperature coefficient by less than 1% on the full panel (LRI interannual variability 0.2771 to 0.2779, +0.3%; asthma 0.8441 to 0.8403, −0.5%). PCV3 coverage itself is null for LRI over the full period (β = 0.0001, p = 0.581) and inversely associated in the 2008–2023 window (β = −0.0003, p = 0.0005). In the restricted sample the LRI mean-temperature and interannual-variability coefficients collapse to null (p = 0.80 and p = 0.26), consistent with the detrending sensitivity: the headline associations are carried by pre-2008 trend variation and are not confounded by staggered vaccine roll-out.*

**Supplementary Table S9. Between/within climate-zone decomposition of the quadratic temperature–mortality U-shape underlying the childhood LRI minimum mortality temperature (MMT), 171 countries, 1990–2023.**

| Model | b1 | b2 (SE) | p (b2) | Implied MMT (°C) | N |
|---|---|---|---|---|---|
| **Panel models (Driscoll–Kraay SE, bandwidth 3, unless noted)** | | | | | |
| M0 baseline: country FE + year FE | −0.3122 | 0.0091 (0.0013) | $1.3\times10^{-11}$ | 17.1 | 5,814 |
| M1 within-zone: country FE + zone × year FE | −0.1170 | 0.0041 (0.0009) | $8.8\times10^{-6}$ | 14.3 | 5,814 |
| M1b within-zone, country-linear detrended | — | 0.0011 (0.0108) | 0.919 | not identified | 5,814 |
| M2 zone-level: zone × year means, zone FE + year FE (zone-clustered SE) | −0.3499 | 0.0139 (0.0045) | 0.003 | 12.6 | 136 |
| **Cross-sectional models on country time means (HC3 SE)** | | | | | |
| M3 between-country, no zone FE | −0.2426 | 0.0079 (0.0068) | 0.240 | 15.3 | 171 |
| M4 between-country, zone FE | 0.0098 | −0.0014 (0.0027) | 0.602 | not identified | 171 |
| **Zone × temperature interaction (single model, country FE + year FE; Wald test of equal b2 across zones: $\chi^2$ = 120.6, p < 0.001)** | | | | | |
| Zone-specific b2: Tropical | — | 0.0230 | $1.3\times10^{-12}$ | 20.3 | 5,814 |
| Zone-specific b2: Subtropical | — | 0.0134 | $1.6\times10^{-6}$ | 17.3 | 5,814 |
| Zone-specific b2: Temperate | — | 0.0004 | 0.785 | not identified | 5,814 |
| Zone-specific b2: Subarctic | — | −0.0040 | 0.006 | not identified | 5,814 |
| Zone-specific b2: Polar | — | 0.0380 | 0.031 | not identified | 5,814 |
| **Variance decomposition of the identifying temperature variation (orthogonal; zone × year FE nest year FE)** | | | | | |
| Zone-level share of identifying temperature variance | | 26.6% | — | — | 5,814 |
| Within-zone share of identifying temperature variance | | 73.4% | — | — | 5,814 |
| Zone-level share of fitted U-signal variance (b1×T + b2×T²) | | 53.1% | — | — | 5,814 |
| Within-zone share of fitted U-signal variance | | 46.9% | — | — | 5,814 |

*Note. All models regress the log LRI mortality rate (ages 0–19) on annual mean temperature and its square, controlling for mean diurnal temperature range, seasonal amplitude, and interannual temperature SD. b1 and b2 are the linear and quadratic coefficients on annual mean temperature; MMT = −b1/(2 × b2) for b2 > 0. M1 replaces year fixed effects with climate-zone × year fixed effects, so the quadratic term is identified only from country deviations from zone–year means; M2 is fitted to the 136 zone × year means of the four named zones; M3–M4 use the 171 country time means with heteroskedasticity-robust (HC3) standard errors. The single-country Polar row (34 country-years) is retained in M0, M1, and the interaction model and excluded from M2. The interaction model's temperate and Polar implied turning points lie far outside the observed temperature range and are reported as not identified.*

*The variance decomposition residualises annual mean temperature (or the fitted U-signal) on country + year fixed effects versus country + zone × year fixed effects; because the fixed-effect spaces are nested, the two components are orthogonal and sum exactly. 26.6% of the identifying temperature variance lies between zones, but this component carries 53.1% of the fitted U-signal variance.*

*Reading: within-zone curvature survives zone × year fixed effects (M1: b2 = 0.0041, $p = 8.8 \times 10^{-6}$) but collapses under detrending (M1b), so it is trend-level like the baseline; the zone-level component is separately U-shaped (M2); the pure cross-section does not pin the minimum on its own (M3). The U-shaped geography is therefore carried by within-zone curvature reinforced by a steeper between-zone trend-level contrast, not by zone aggregation alone.*
